# Change in disease dynamics and health care utilisation in children during COVID 19 in a tertiary care hospital of Eastern India

**DOI:** 10.1101/2021.06.24.21259435

**Authors:** Bandya Sahoo, S Suneeti Kanyari, Deepti Damayanty Pradhan, Sibabratta Patnaik, Manas Ranjan Behera

**Affiliations:** Kalinga Institute of Medical Sciences, Bhubaneswar, India

**Keywords:** COVID-19, Disease dynamics, Health care, Tertiary care hospital

## Abstract

This retrospective, observational study was conducted by collecting data from medical records during COVID 19 pandemic from March 2020 till August 2020. This was compared with the data of 2019 during similar months. The impact of COVID 19 on use of preventive and curative paediatric health care service units like outpatient department, casualty, intensive care and immunization clinic were assessed. Data from 2019 to 2020 were compared using standard parametric and nonparametric tests. There was a significant decline in routine OPD (68%) attendance during the COVID 19 period as compared to pre-COVID period. Paediatric ward admissions and PICU admissions were decreased by 55% and 42% respectively. We also observed a significant 43% decline in the number of children attending immunization clinic in the year 2020. The fear of COVID 19 pandemic and the measures taken to control the pandemic has affected the health seeking behaviour of patients. This evaluation of trends in healthcare use may help in planning the delivery of healthcare service delivery in future.

## Introduction

The direct and indirect impact of SARS COVID 19 on the health of children was unprecedented. Effect of the pandemic on epidemiology of other diseases was unanticipated. The impact of the disease has been far beyond acute and chronic diseases. The two months long lockdown in 2020 in our country had consequent effect on social interactions amongst children and also affected their mental health. The lockdown along with the fear of the disease made access to various health services difficult. Because of restriction of movement and social distancing the transmission of infectious diseases decreased. COVID 19 adversely affected both health care services for diseases and disease preventive services i.e. immunization. ^[1]^

Social isolation and decreased outdoor visits reduced the risk of infection and infectious diseases. Lockdown imposed minimal travel and decreased mobility and thereby decreased the incidence of injuries and road traffic accidents. ^[2]^ The change in lifestyle, with parents spending more time at home, resulted in lesser number of in house accidents and accidental poisoning. Moreover, the decreased likelihood of direct infection with SARS-Co V-2 in children ^[3, 4]^ as compared to adults reduced the healthcare visits related to the disease itself. These could be some of the reasons for a modified pattern of healthcare utilization and reduction in the number of visits to hospital during the SARS-CoV-2 outbreak. ^[5, 6]^

The literature in this regard is constantly evolving and varies according to the geography and demography of the patients. Therefore this study was conducted to compare the changing pattern of disease dynamics and the use of healthcare system before and after the SARS-CoV2 outbreak in a tertiary care hospital.

## Materials and methods

This retrospective, observational study was conducted in the department of Pediatrics of Kalinga institute of Medical Sciences, a tertiary care hospital after obtaining clearance from institutional ethics committee. Medical records of COVID 19 pandemic period, were reviewed i.e. March 2020 till August 2020. This was compared with the Pediatric OPD and Emergency Department attendance during pre-COVID 19 period i.e. March 2019 to August 2019. All patients who attended the Pediatric OPD, immunization clinic and emergency were included in the study. Patient profile included; the demographic and clinical characteristics along with outcome. The clinical features and diagnosis were grouped into 2 broad categories. Patients who used preventive service (immunization) and those who used curative service (OPD and emergency). Those who attended curative service were grouped into a) Acute respiratory illness b) Acute infectious diseases c) Trauma 4) Poisoning 5) Acute surgical conditions 6) Chronic diseases and 7) other diseases. Diagnosis and discharge were coded as per International Statistical Classification of Diseases and Related Health Problems (ICD 10-AM). Total number of patients visiting the hospital daily and the admission rate was noted. Admission rate was calculated as the number of admissions from the casualty and OPD relative to the total number of visits.

### Statistical analysis

The data including demographic characteristics of all patients and their disposition either to ward, pediatric intensive care unit (PICU) or not admitted was collected in a proforma. The continuous variables were presented as mean and standard deviation, while the categorical variables as proportions and percentages. Categorical variables were analyzed using chi-square test. Data from 2019 to 2020 were compared using standard parametric and nonparametric descriptive tests. The variables having normal distribution were analysed using independent sample t-test and the variables having non-normal distribution were analysed using Mann-whitney u test. Statistical analyses were performed using SPSS for Windows, version 21.0. P value of < 0.05 was considered as statistically significant.

## Results

A total of 20,018 children visited the hospital during pre-COVID (March 2019 to August 2019) as compared to 6,377 in COVID (March 2020 to August 2020) period. The number of children visiting for immunization was 2,011 and number of patients visiting the emergency department and OPD were 1,939 and 4,438 respectively in the COVID period. There was a significant decrease in the mean number of monthly patients visits (3336±555) during COVID as compared to the pre-COVID in 2019 (1063 ±833, p<0.001).

There was a significant decline in routine OPD and casualty attendance as well as the total number of hospital admissions during the COVID period as compared to pre-COVID period as shown in Table 1. Routine OPD attendance was reduced by 68%. Likewise the paediatrics casualty attendance reduced from a monthly average of 1045 in the year 2019 to 323 in 2020: a significant 69% decline. Paediatric ward admissions and PICU admissions were decreased by 55% and 42% respectively. There was also a significant 43% decline in the number of children attending immunization clinic in the year 2020.

**Table 1:**
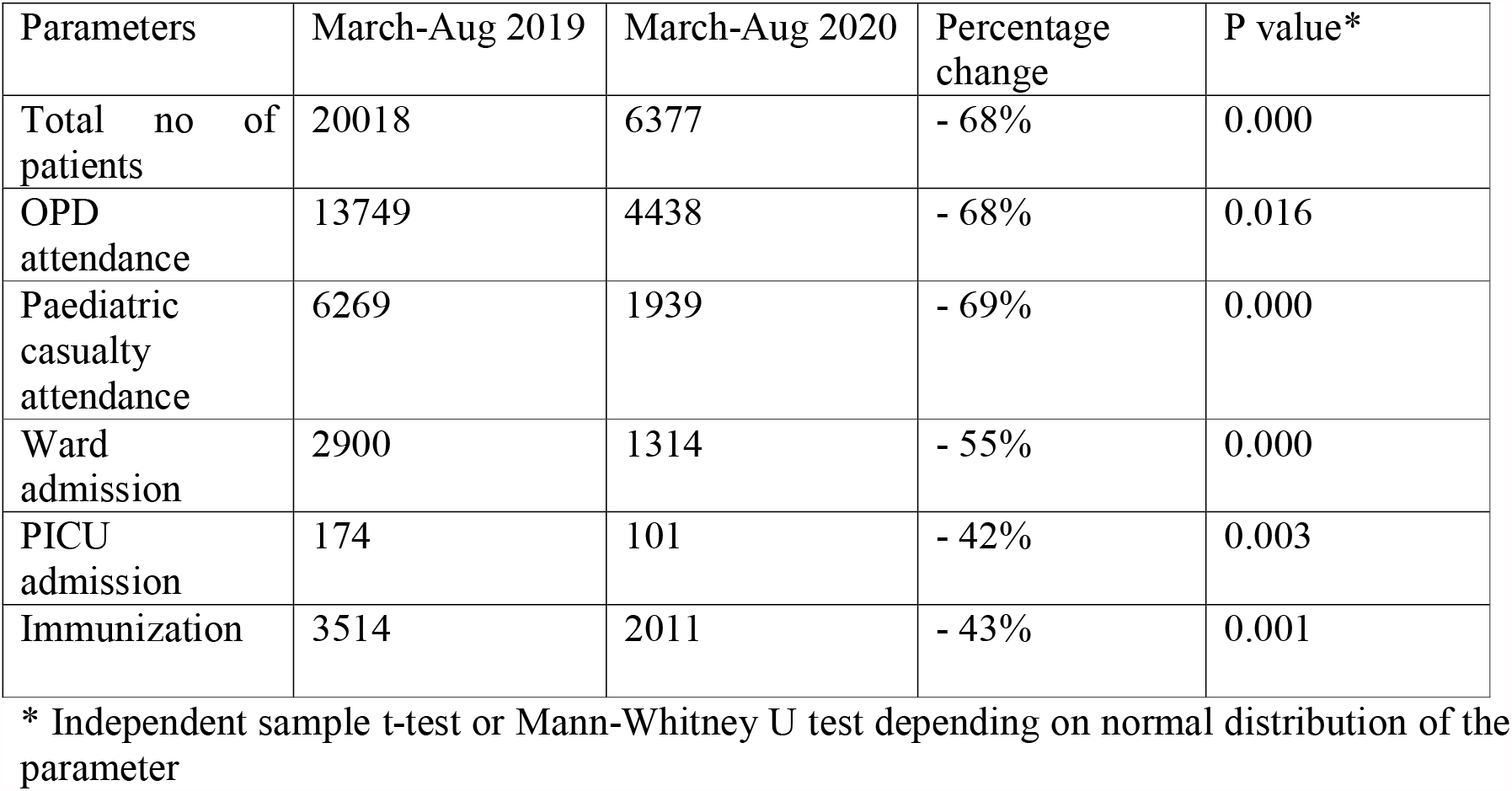
Impact of COVID-19 on hospital attendance and admissions.

There was a significant decline of 30% in the number of children being admitted to NICU during COVID period. The mean number of births decreased from 163 during pre-COVID period to 104 during COVID period – a reduction of 36%. The decline in number of deliveries, 24% decline in normal delivery and 45% decline in delivery conducted by caesarean section, was found to be statistically significant. (Table 2)

**Table 2:**
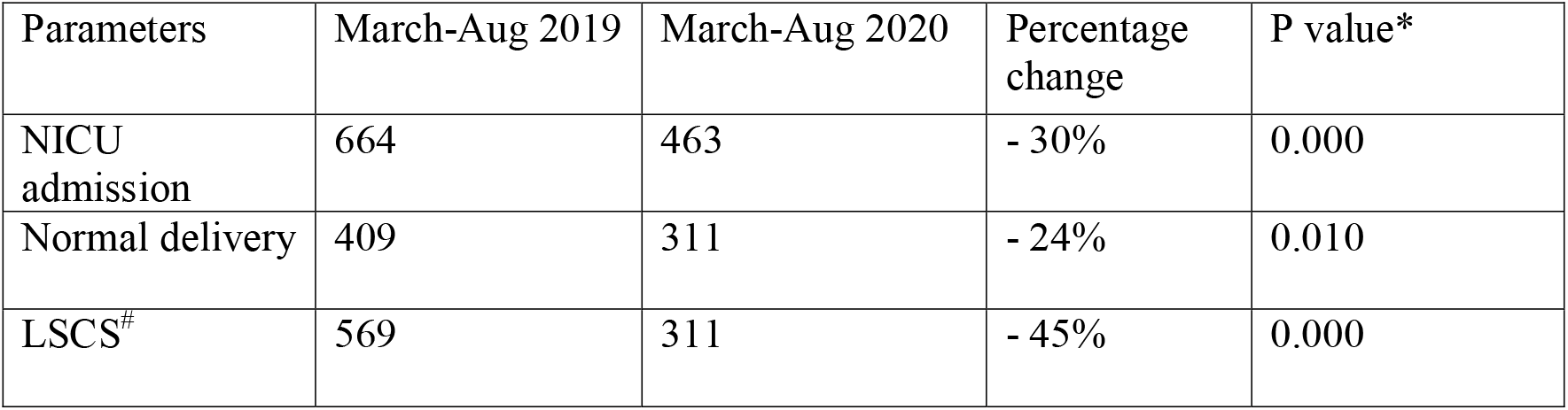
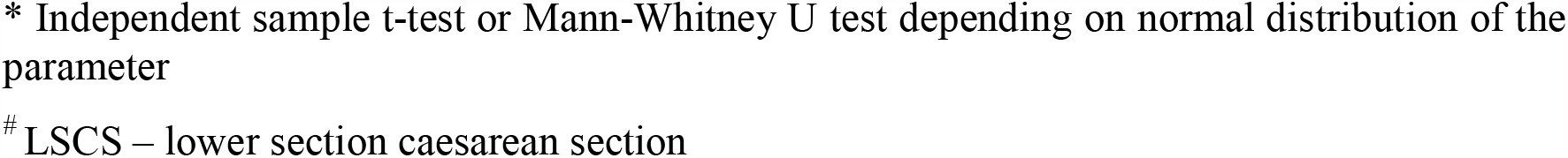
Impact of COVID-19 on neonatal and maternal health services.

Reductions in paediatric ward admissions were noticed uniformly across all disease categories as shown in Table 3. Patients who presented with disease categories of acute respiratory illness, acute infectious diseases, trauma, poisoning and other diseases showed a statistically significant difference between pre-COVID and COVID period. However the difference was not significant in case of acute surgical conditions and chronic diseases.

**Table 3.**
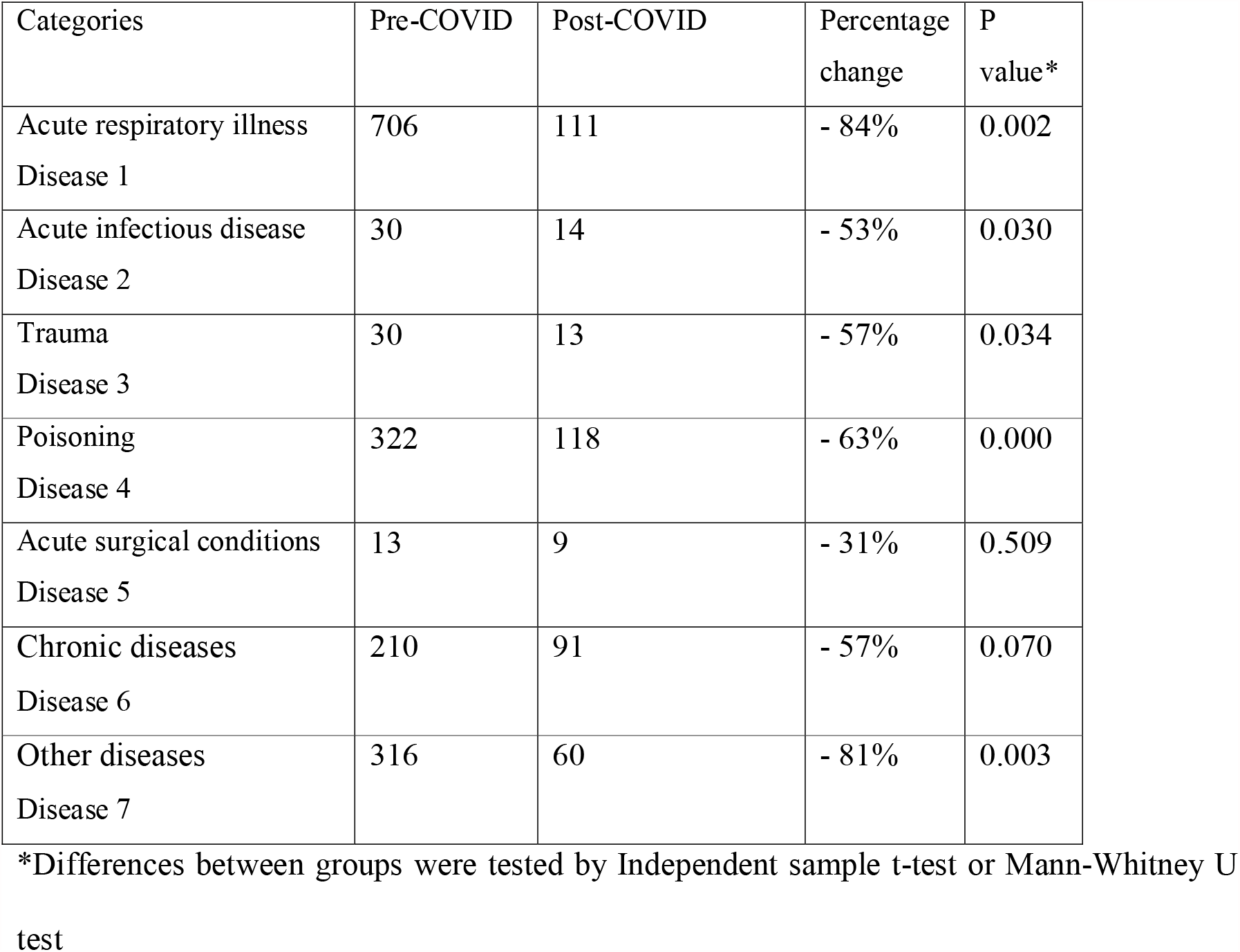
Comparison of total number of hospitalizations according to disease categories in the same time period.

## Discussion

Children have been largely spared from the direct impact of COVID pandemic, although a minority of them were affected with SARS CoV2 related hyper-inflammatory syndrome. ^[7]^Hence the need for emergency medicine care was less as compared to adults. ^[8]^ But delayed presentation of acute and critically ill children increased which could be due to unwillingness in seeking timely medical attention because of fear of COVID. Data from pediatric providers have demonstrated that approximately 1 in 3 presentations for emergency medical care was perceived to be delayed. ^[9]^

This study reports the effect of COVID-19 on the number and type of presentation of children. There was a significant decline in the total number of pediatric attendance by 68% and a substantial decrease in the rate of hospitalizations during COVID pandemic. Similar decrease in number of visits during COVID pandemic has been reported in other studies also.^[7]^ Isba et al reported a fall in attendance in UK by 33.8% ^[10]^, with a decrease in attendance by 38.8% and 50% in Argentina and Ireland respectively. ^[11, 12]^ Sonadi et al documented a fall in total pediatric attendance by 54%, and emergency department attendance reduced by 43% in India. ^[13]^ The reason for this could be restriction of movement imposed by lockdown and denied access to health care facilities due to fear of COVID -19 transmission. The reduction in hospital visits was found in all disease categories. There was a marked decrease in visits in children aged 1-5 years. A similar finding has been recorded by Sonadi et al. The trauma patients reduced from 30 in pre-COVID to 13 in COVID period. This could be due to lockdown which lead to children staying at home and thereby decreased road traffic accidents and elimination of injuries at school. Jang et al also reported a decrease in trauma cases by 3.9% although proportion of trauma patients markedly increased by 20.5% in infants and young children ^[14]^ which they attributed to possibility of inability to control posture or judge a situation. Children diagnosed with chronic/underlying diseases was not significantly (P 0.07) affected during the COVID-19 pandemic. Liguoro et al reported a similar finding in their study from Italy. ^[15]^ On the contrary, there was a significant (P -0.005) reduction in children with respiratory illnesses which could be due to a segregation of patients into Flu OPD. Also less movement leading to less transmission.

Studies have estimated a significant impact of the pandemic also on maternal and neonatal health services. This study shows a significant decrease by 30% in NICU admission. Liu et al reported a similar finding in their study. ^[16]^ Our study shows a significant decline of 24% and 45% in normal and caesarean section deliveries respectively. A similar finding has been reported in Nepal by Bhogadi et al. ^[17]^ Thus, the challenges faced by the pediatric intensive care unit included not only the different disease pattern but also delayed presentation of critically sick children. ^[18]^

## Conclusion

The changing patterns of healthcare use during COVID 19 pandemic could be due to decreased need for emergency care and decreased use of emergency medical care. This could be due the fear of COVID pandemic leading to change in the health seeking behaviour of patients. A significant decrease in acute respiratory, infectious illness and trauma could be attributed to the lockdown and social distancing. This evaluation of trends in healthcare use may help in planning the delivery of healthcare service delivery in future.

### What’s already known

COVID 19 lead to decreased utilization of both curative and preventive health services in children

### What the study adds

The possible sequelae of the decrease in healthcare utilization in the form of delayed diagnosis of developmental diseases and re-emergence of vaccine-preventable diseases, should be looked for in future.

## Data Availability

YES

